# Innate immune genes distinguish the immune microenvironment of early onset colorectal cancer

**DOI:** 10.1101/2020.06.26.20141143

**Authors:** Ivy H. Gardner, Ragavan Siddharthan, Katherine Watson, Elizabeth Dewey, Rebecca Ruhl, Xiangnan Guan, Zheng Xia, Liana V. Tsikitis, Sudarshan Anand

## Abstract

Despite a decrease in the incidence of colorectal cancer (CRC) over the last 40 years, the incidence of CRC in people under 50 years old is increasing around the globe. Early onset (≤50 years old) and late onset (≥65 years old) CRC appear to have differences in their clinicopathological and genetic features, but it is unclear if there are differences in the tumor microenvironment. We hypothesized that the immune microenvironment of early onset CRC is distinct from late onset CRC and promotes tumor progression. We used Nanostring immune profiling to analyze mRNA expression of immune genes in FFPE surgical specimens from patients with early (N=40) and late onset (N=39) CRC. We found three genes, SAA1, C7, and CFD, have increased expression in early onset colorectal cancer and distinct immune signatures based on the tumor location. After adjusting for clinicopathological features, increased expression of CFD and SAA1 were associated with worse progression free survival and increased expression of C7 was associated with worse overall survival. Our data demonstrate that the immune microenvironment in early onset CRC is unique, location dependent, and associated with worse outcomes.

## Introduction

Colorectal cancer (CRC) is the second leading cause of cancer-related death in the United States (1). Over the last 40 years, there has been an overall decrease in the incidence of CRC (1, 2) attributed to current screening guidelines (1, 2). Despite this decrease, there has been an increase in the incidence of CRC in young adults under the age of 50 over the same time period (2, 3). In particular, between 1980 and 2010, the incidence of rectal cancer in people aged 20-39 has quadrupled and it appears that the overall increase in early onset CRC (≤50 years old) is largely driven by an increase in rectal cancer in young patients (4). Based on these trends it is predicted that by the year 2030, the incidence of colon cancer will increase by 90% and rectal cancer will increase by 124% in people aged 20-34 (2). In addition, the rates of advanced disease manifested as localized, regional, and distant CRC have increased in patients 40-49 years old (5). This indicates that the increase in incidence is real and does not represent a shift in the age of diagnosis due to earlier detection with screening (5).

The etiology of the rise in early onset CRC is unclear, but it appears to be global (6–8). The proportion of CRC cases occurring in young patients has risen more in the highest income quartile, urban population, and Western states in the United States (3, 9). This rise in early onset CRC does not correlate with patterns of obesity or heavy alcohol use (9). Young patients with CRC are more likely to present with regional or metastatic disease, high grade, mucinous, and signet ring tumors (3, 7, 8, 10–16). Early onset is more commonly located in the rectum and left colon compared to late onset CRC (7, 14–16). Patients with early onset CRC appear to have similar or improved stage-specific and overall survival (7, 10, 11, 13, 16).

The majority of early onset CRC are sporadic with only 16% of early onset CRC cases having genetic mutations (17). About 10-21% of early onset CRCs are mismatch repair deficient with 6-8% having Lynch Syndrome and these cancers are usually right sided (17–19). In addition to distinct clinicopathological features, early onset CRC appears to have molecular differences to late onset CRC. Compared to late onset CRC, early onset CRC has lower rates of BRAF mutations and methylator phenotype (20, 21). Early onset microsatellite stable CRC has more mutations in TP53 and CTNNB1 compared to late onset microsatellite stable CRC (22). Several genes and key pathways in early onset CRC have been identified and include PEG10, the Wnt/beta catenin, MAP kinase, growth factor signaling, PI3KT-AKT, and TNFR1 pathways (20, 23–25).

The immune microenvironment in CRC is a key player in disease progression, therapy response, and overall survival (26–30). To our knowledge the role of the immune microenvironment in early onset CRC has not been well-studied. We set out to characterize the differences between the immune microenvironment in early onset (≤50 years old) and late onset (≥65 years old) CRC in a well-annotated cohort of patients at our tertiary cancer center.

## Combined Results and Discussion

Our institutional cohort consisted of 40 early onset and 39 late onset CRC patients. A higher percentage of the early onset CRC group had metastatic disease and therefore underwent more neoadjuvant and adjuvant treatments (Figure 1A). Additionally, our early onset CRC group had less microsatellite instability compared to the late onset group (Figure 1B). We performed gene expression profiling for 770 immune response genes using the NanoString Immune panel from RNA extracted from FFPE tissues. We found 3 immune genes significantly (adjusted p<0.1) upregulated in early onset CRC (SAA1, C7, CFD) and 7 genes upregulated in late onset CRC (CXCL3, CCL18, IL8, DMBT1, CEACAM6, CXCL5, CXCL10) (Figure 2A, 2E, and Supplemental Figure 1). Right colon, left colon, and rectal cancer have differentially expressed immune genes between early and late onset groups (Figure 2B-D). The increased expression of SAA1, C7, and CFD in early onset CRC is largely driven by an increase in rectal cancer, although C7 was also increased in early onset right colon cancers. We identified 10 genes of interest that were differentially expressed between early and late onset CRC in the whole cohort and at least one location specific group (SAA1, C7, CFD, CXCL12, DMBT1, CEACAM6, CCL18, IL8, CXCL3, CXCL10).

**Figure 1:**
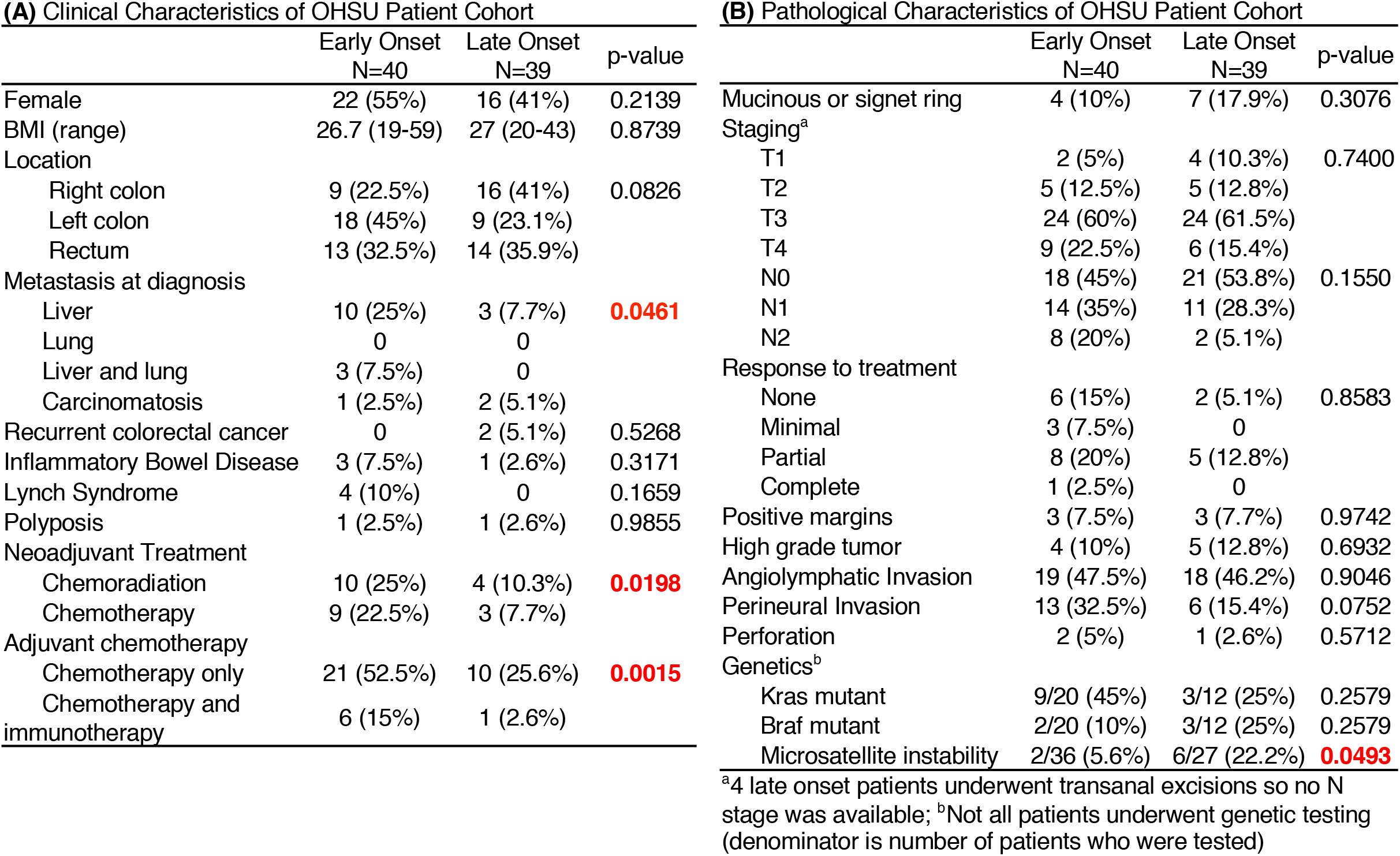
Clinical and pathological characteristics of institutional cohort. Descriptive statistics of (A) clinical and (B) pathological characteristics of early (N=40) and late (N=39) onset colorectal cancer cohorts. Early and late onset patient cohorts were compared using a two-tailed unpaired T-test for continuous variables and chi-squared test was used for categorical data.

**Figure 2:**
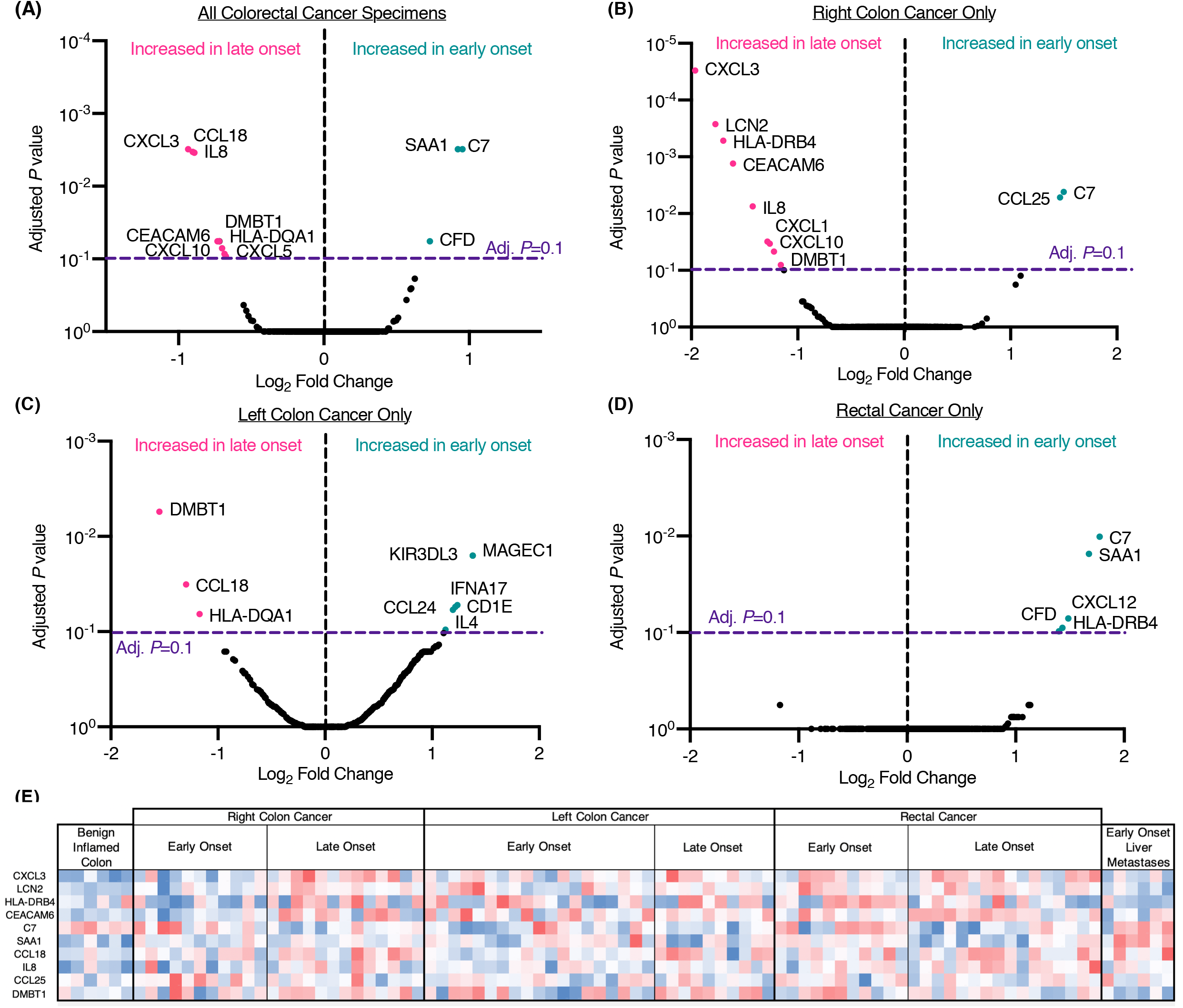
Immune microenvironment differences with age in colorectal cancer. Volcano plots depicts log 2 fold change in gene expression vs adjusted P-values of early vs late onset colorectal cancer patients. Nanostring Immune profiling panel (770 genes, human cancer immune panel) was used to generate expression profiles from RNA extracted from FFPE tissues slides. Raw counts were normalized to the best 10 housekeeping genes. The nSolver Advanced Analysis module was used to derive differential expression signatures according to Nanostring recommendations. Adjusted P-values were obtained with a Benjamini-Hochberg post-hoc correction. (A) Early (N=40) vs late onset (N=39) in all colorectal cancer specimens. (B) Right colon cancer specimens only (early N=10, late N=14). (C) Left colon cancer specimens only (early N=19, late N=10). (D) Rectal cancer specimens only (early N=11, late N=15). (E) Heatmap of the top 10 most differentially expressed immune genes with blue representing low relative gene expression and red representing high relative gene expression compared to other specimens.

We validated the increased expression of one gene (CXCL3) in late onset colorectal cancer and another gene (SAA1) in early onset rectal cancer using qRT-PCR assays from the same patient samples (Figure 3A-B). We also found that SAA1 gene expression was higher in a group of liver metastases, but the significance of this finding is unclear because we did not have access to non-tumor liver samples within our colorectal cancer registry to compare to. Additionally, SAA1 expression was lower in a group non-tumor samples compared to early onset rectal cancer. We analyzed a publicly available dataset (GSE87211) of gene expression microarray profiles from rectal cancer (N=105) and matched adjacent mucosa specimens (N=84) to further validate our findings (31). In this rectal cancer dataset, we found a difference in CXCL3 expression between tumor and mucosa in both groups, but no difference in gene expression between early and late onset rectal tumor specimens (Figure 3C). This is congruent with our findings since CXCL3 is not differentially expressed in rectal cancer in our cohort. We found that SAA1 levels were increased in the early onset tumors compared to late onset tumors, but there was no difference in expression between age groups in the adjacent mucosa (Figure 3D). This suggests that the upregulation of this gene may not simply be a product of immune aging, but more specifically due to the tumor immune microenvironment in early onset CRC.

**Figure 3:**
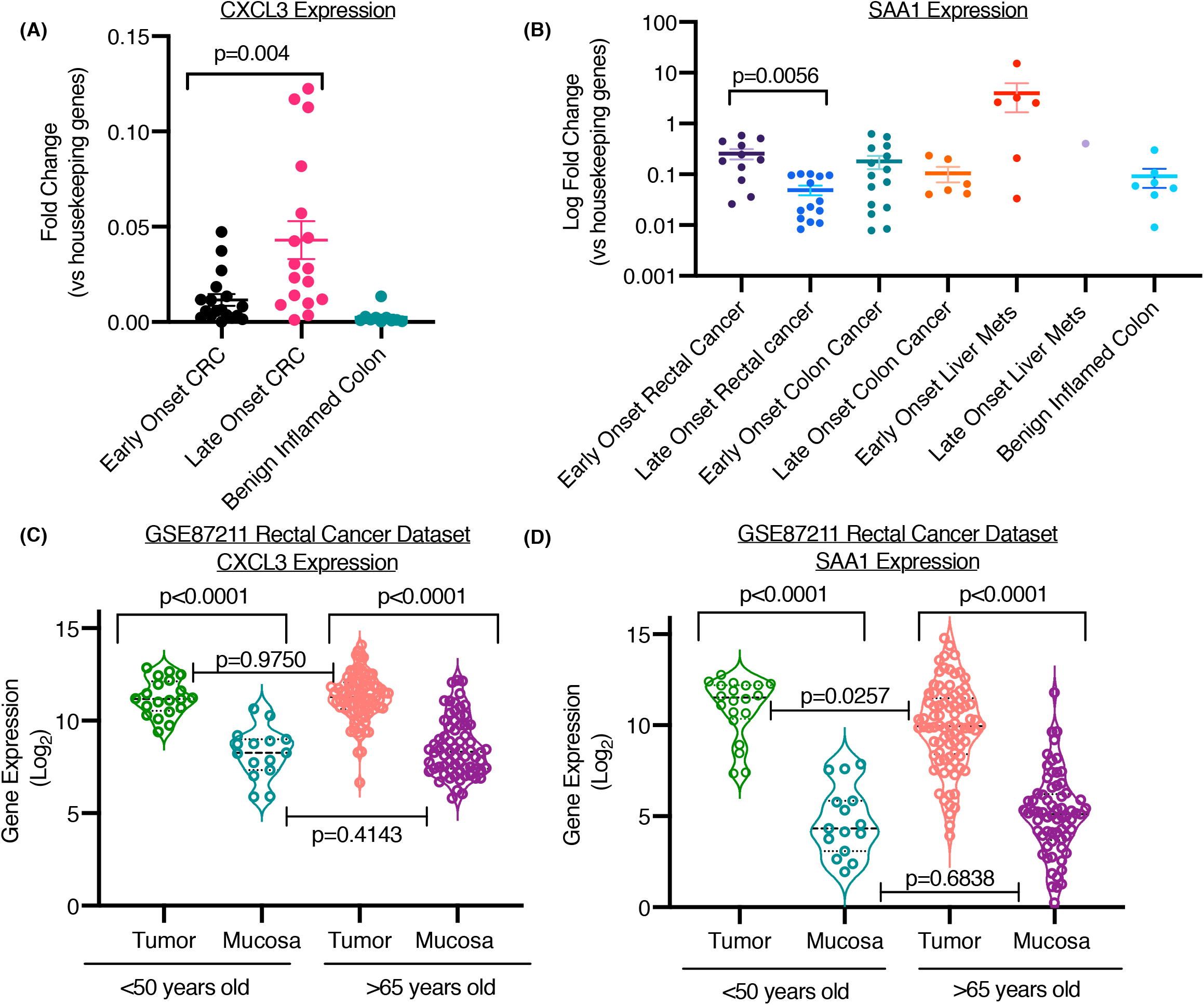
Validation of Nanostring data with independent qRT-PCR assays and independent datasets. (A) CXCL3 and (B) SAA1 expression of individual tumor RNA samples from FFPE slides measured using Taqman probes in a qRT-PCR assay. Change in gene expression (2-DCt) are depicted as fold change over 2 housekeeping genes. Violin Plots depict (C) CXCL3 and (D).SAA1 gene expression from a publicly available Rectal Cancer dataset GSE87211. Values are log gene expression measured via microarrays in tumors and matched adjacent mucosal samples. P-values are from a two-tailed unpaired T-test with Welch’s correction.

We found several associations between the expression of our genes of interest and the clinical characteristics, pathological features, and outcomes of our patient population. Increased expression of C7 is associated with neoadjuvant treatment, right colon location, metastatic disease, angiolymphatic and perineural invasion (Supplemental Figure 2). Elevated CFD is only associated with neoadjuvant treatment and SAA1 does not appear to be associated with any clinical or pathological features (Supplemental Figure 2). After adjusting for age, metastatic disease, angiolymphatic invasion, perineural invasion, tumor grade, and neoadjuvant treatment, we found increased expression of SAA1 and CFD are associated with worse progression free survival (Figure 4A) and increased expression of C7 is associated with worse overall survival (Figure 4B). High expression of C7 and CFD was also associated with worse overall survival in The Cancer Genome Atlas Colon Adenocarcinoma (COAD) cancer cohort (Supplemental Figure 3). The combination of SAA1, C7, and CFD as an immune signature may have a role in predicting poor progression free and overall survival.

**Figure 4:**
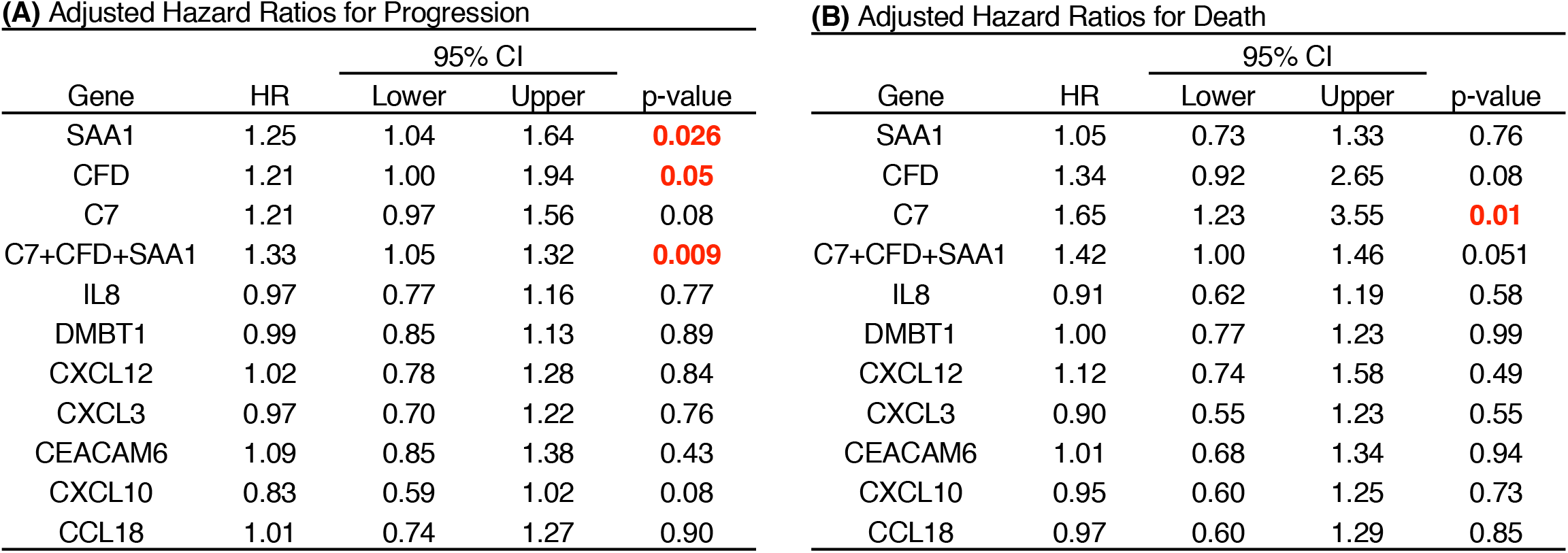
Adjusted Hazard Ratios. Hazard ratios for (A) progression and (B) death were calculated using multivariable Cox Proportional Hazards and are adjusted for age, metastasis at diagnosis, angiolymphatic invasion, perineural invasion, neoadjuvant therapy, and tumor grade. Hazard ratios are per half of a standard deviation from the mean of gene expression.

We found that elements of complement (C7, CFD) and SAA1 have increased expression in early onset CRC and are associated with worse outcomes. Analysis of gene set enrichment signatures from our profiles indicate complement and B-cell function is the most enriched pathway in early onset CRC (Supplementary Figures 4). C7 and CFD play a role in the terminal and alternative pathways of complement, but components of complement also have a non-canonical role that modulates the immune system and plays a role in the tumor immune microenvironment (32). Complement can play a pro-tumorigenic or anti-tumor effect in the tumor immune microenvironment and it is unclear what role C7 and CFD play in CRC (32). Complement activation in intestinal epithelial cells has been shown to be associated with inflammation; therefore the increase in C7 and CFD in early onset CRC may be related to an underlying inflammatory process (33). Similar to complement, SAA1 has also been shown to be overexpressed in patients with active inflammatory bowel disease (34, 35). SAA1 is an acute phase reactant produced by the liver, but is also present in multiple cancers and can be used as a biomarker for disease burden (36–38). SAA1 has been shown to direct pathogenic Th17 cell differentiation (34, 35). It is unclear if the increased expression of CFD, C7, and SAA1 in early onset CRC is the result of underlying immune dysregulation leading to a tumor-permissive microenvironment or an inappropriate immunological response to malignancy.

One of the hallmarks of immune aging is the phenomenon of “inflammaging” which is characterized by low levels of persistent inflammation both systemically and within the tissue microenvironment (39). Although the majority of research on immune system aging has been on adaptive immunity, the phenomenon of inflammaging is now largely attributed to innate immune system dysfunction (39). Immune aging appears to be driven by a complex interaction between genetic and environmental factors and appears to exist along a trajectory towards an older immune system with people advancing across the trajectory at different speeds (40). Our findings paradoxically suggest that early onset CRC is associated with an inflammatory phenotype which is what would be expected in an older population. It may be that early onset CRC is associated with expedited inflammaging that leads to a tumor-permissive microenvironment. We found that SAA1, an acute phase protein, is elevated in early onset CRC which is akin to the elevation of another acute phase protein, C-reactive protein, in inflammaging. Studies have shown that older individuals have reduced microbiota diversity, decreased density of beneficial bacteria, and weakened integrity of the intestinal barrier (41). It is hypothesized that these factors allow leakage of proinflammatory microbial products into the systemic circulation and peripheral tissues which leads to the chronic inflammation associated with aging (41, 42). The microbiome and dysregulation of the intestinal barrier may play a role in the etiology of early onset CRC.

Our findings point to the innate immune system, specifically complement, as a component of the immune microenvironment that distinguishes early onset CRC. Although complement has been shown to have both protumor and antitumor effects, therapeutics targeting complement are currently being investigated. The majority of studies on complement inhibition in the treatment of cancer have been done in mice. Both C3a and C5a receptors may be potential targets for cancer immunotherapy and the inhibition of C5a in mice has been shown to decrease metastasis in colon cancer (32, 43, 44). Additionally, inhibition of the C5a receptor in mice reduces cancer promoting inflammation that results from consumption of a high-fat diet (45). Of note, there is currently a phase I clinical trial for a C5aR1 monoclonal antibody in combination with durvalumab in advanced solid tumors (32). Our study suggests that early onset CRC may benefit from the addition of a complement inhibitor in addition to the standard use of chemotherapy and immunotherapy. Another possible therapeutic route in early onset CRC is manipulation of the microbiome. Mouse studies have shown that the microbiome appears to play a role in the effectiveness of chemotherapy and immunotherapy, including immune checkpoint inhibitors (46, 47). Manipulation of the microbiome by diet alterations, probiotics, antibiotics, or fecal transplant are being investigated as potential avenues to improve responsiveness to cancer treatments (47). Our findings suggest an inflammatory tumor promoting phenotype in early onset CRC and these treatments may be particularly useful in combating this immune microenvironment.

Our data demonstrate that the immune microenvironment is different between early and late onset CRC and these differences do not appear to solely be the result of age-related immune changes. Additionally, differences in the immune microenvironment between early and late onset CRC are location specific. C7, CFD, and SAA1 are increased in early onset colorectal cancer and are associated with worse outcomes. These immune genes are associated with intestinal inflammation, suggesting that the immune microenvironment in early onset CRC may be pro-inflammatory and tumor permissive as a result of immune dysregulation.

## Methods

The Oregon Colorectal Cancer Registry was queried to find patients with CRC who underwent surgical resection between January 2008 and October 2019 and were either under 50 or over 65 years old at the time of their diagnosis. In addition, six patients who underwent sigmoid colectomy for diverticulitis were identified to serve as a control group. The electronic medical record was used to obtain patient demographics, neoadjuvant treatment, pathology, genetic mutational status, adjuvant treatment, and outcomes.

Formalin-fixed paraffin-embedded (FFPE) slides were reviewed by pathology and only used if at least 70% of the tissue was tumor. The FFPE blocks were sectioned into 5µm-thick slides of tissue for use. Hematoxylin and Eosin stained slides were prepared for each sample to identify the tumor and were used as guides to sharply dissect the tumor from the unstained slides to be used for RNA extraction. RNA was isolated using the RNeasy FFPE kit (Qiagen). RNA concentration and purity was confirmed with the NanoDrop spectrophotometer (ThermoFisher Scientific).

NanoString immune profiling was performed using the human PanCancer Immune panel (cat#115000132) and run on the NanoString nCounter SPRINT profiler. We chose to use the NanoString platform since this platform had better tolerance for moderate RNA integrity scores typical of archival FFPE specimens. 100ng of RNA extracted from the FFPE tissue was used per the manufacturer’s instructions.

Quantitative-PCR analysis was performed to validate expression of two genes identified as differentially expressed based on the NanoString analysis using TaqMan Gene Expression Assays (Applied Biosystems, Cat# 4448892, SAA1 Hs00761940_s1 and CXCL3 Hs00171061_m1) and the ViiA7 qRT-PCR machine (Applied Biosystems). Two housekeeping genes, GAPDH and ACTB (Applied Biosystems, Cat#433182 (Hs02758991_g1, Hs001060665_g1)), were used for all samples. Gene expression was reported as the change in the cycle threshold (−2^ΔCt^, normalized to housekeeping genes).

### Statistics

Early and late onset patient cohorts were compared using a two-tailed unpaired T-test for continuous variables and chi-squared test was used for categorical data. Raw NanoString immune gene expression counts were normalized to the best 10 housekeeping genes. The nSolver Advanced Analysis module was used to derive differential expression signatures according to NanoString recommendations. Adjusted P-values were obtained with a Benjamini-Hochberg post-hoc correction. The genes with an adjusted p value of less than 0.2 in the entire colorectal cancer cohort and at least one location specific cohort (right colon, left colon, or rectum) were designated genes of interest. This short list of 10 genes was then used for further analysis. Hazard ratios were calculated using multivariable Cox Proportional Hazards, and all estimates were adjusted for age, metastasis at diagnosis, neoadjuvant therapy, angiolymphatic invasion or perineural invasion, and tumor grade. The PH assumption was verified using time-dependent covariates. Comparisons between gene expression for qPCR results was done with a two-tailed unpaired T-test with Welch’s correction. Statistical analysis was performed using GraphPad Prism version 8.4.2 (GraphPad Software Inc., San Diego, CA), and JMP 14.3 (SAS Inc. Cary, NC).

### Public Datasets

The GSE87211 rectal cancer gene expression microarray dataset was used to compare expression of our genes of interest in rectal tumor specimens and matched adjacent mucosa (31). This dataset was downloaded and divided into early (<50 years old) and late onset (>65 years old) groups. Tumor and mucosa specimens were compared between groups using a two-tailed unpaired T-test with Welch’s corrections.

To further examine the correlation of the up-regulated genes with the clinical survival outcomes in the independent and large datasets, we investigated their expressions in bulk RNA-seq data from The Cancer Genome Atlas (TCGA). The processed gene expression data (fragments per kilobase per million fragments) as well as clinical data for primary colon adenocarcinoma (COAD) solid tumors were downloaded using the Bioconductor *TCGAbiolinks* package (version 2.14.0). Patients with overall survival less than 10 days were excluded. The remaining COAD tumor samples were stratified into the high and low groups based on the top quantile and bottom quantile of the expression of each individual gene. The overall survival curves of these two groups of patients were estimated by the Kaplan-Meier method with statistical significance calculated by the log-rank test.

### Study Approval

This study was approved by the OHSU Institutional Review Board (study #00021278). Only patients who had undergone informed consent to be included in the Oregon Colorectal Cancer Registry were included in this study.

## Author Contributions

IG-planning experiments, conducting experiments, analyzing data, writing manuscript

RS-planning experiments, conducting experiments, analyzing data, editing manuscript

KW-planning experiments, conducting experiments, writing manuscript

ED-analyzing data, editing manuscript

RR-planning experiments, conducting experiments, analyzing data, editing manuscript

XG-analyzing data, editing manuscript

ZX-analyzing data, editing manuscript

LVT-planning experiments, writing manuscript

SA-planning experiments, analyzing data, writing manuscript

## Data Availability

Data that is not included in the supplementary material is available upon request from the corresponding author.

## Acknowledgements

This work was supported by funding from NHLBI to S.A. (R01 HL137779 and R01 HL143803). We thank members of the Anand lab for useful discussions.

---

**Supplemental Figure 1:**
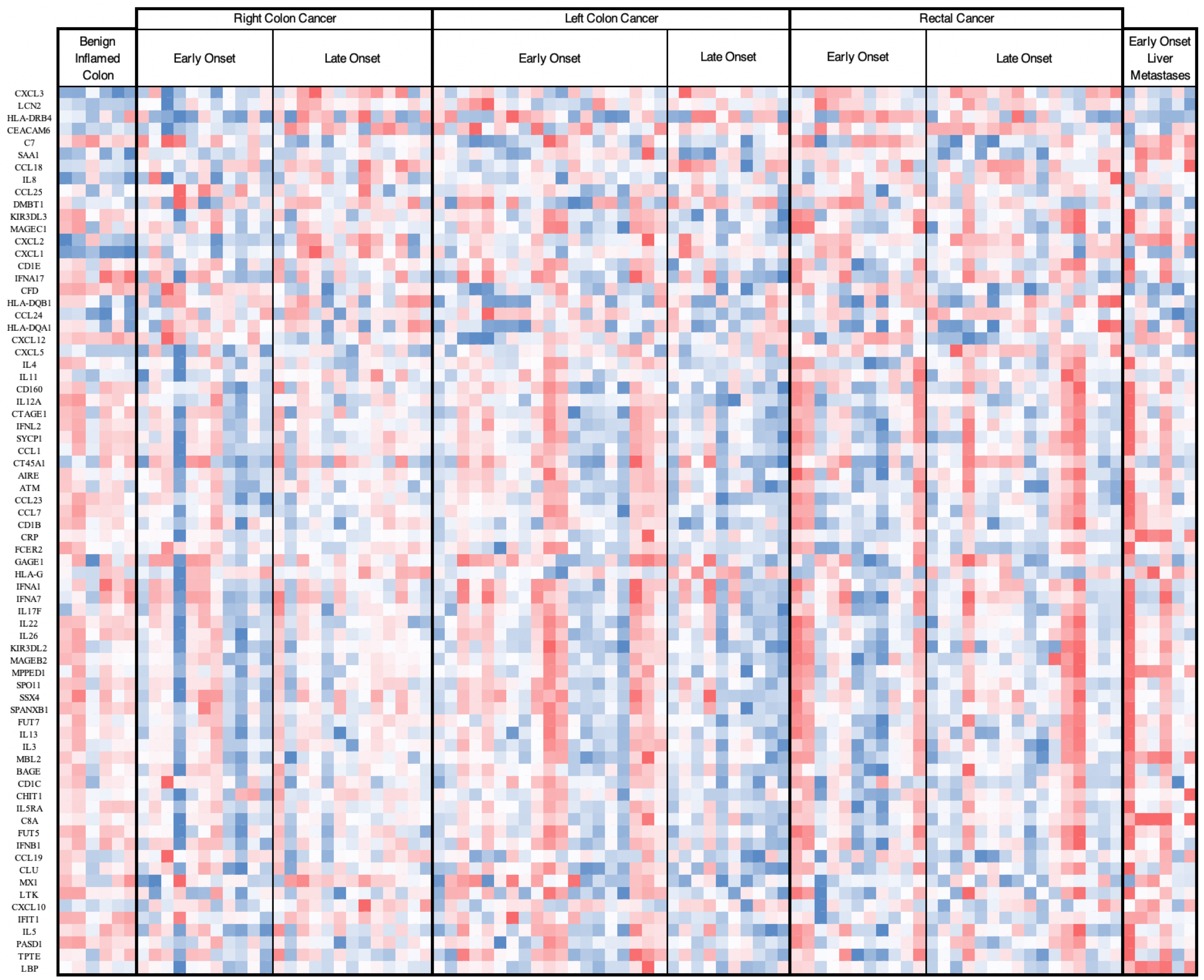
Heatmap of differentially expressed immune genes between early and late onset colorectal cancer. Nanostring Immune profiling panel (770 genes, human immune panel) was used to generate expression profiles from RNA extracted from FFPE tissues slides. Raw counts were normalized to the best 10 housekeeping genes. The nSolver Advanced Analysis module was used to derive differential expression signatures according to Nanostring recommendations. Adjusted P-values were obtained with a Benjamini-Hochberg post-hoc correction. Genes with an adjusted p value >0.2 in all colorectal cancer, right colon cancer, left colon cancer, and rectal cancer specimens are included in the heatmap. The genes are ordered by p values with lowest p values at the top. Blue represents low relative gene expression and red represents high relative gene expression compared to other specimens.

**Supplemental Figure 2:**
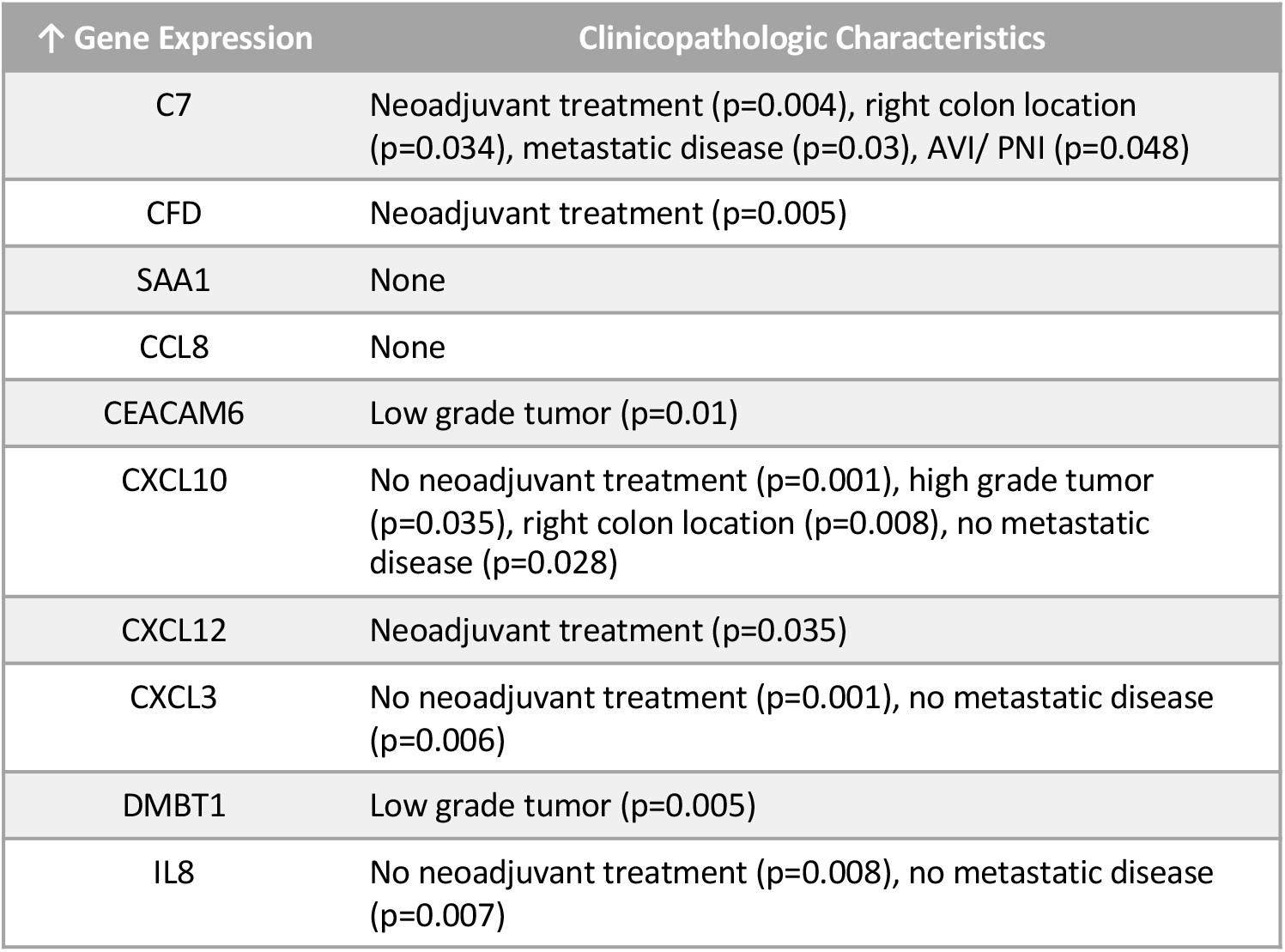
Associations between immune gene expression and clinicopathologic characteristics. Associations between immune gene expression and neoadjuvant treatment, tumor grade, tumor location, metastatic disease, angiolymphatic invasion (AVI), and perineural invasion (PNI). Other clinicopathological features were not significantly associated with gene expression.

**Supplemental Figure 5:**
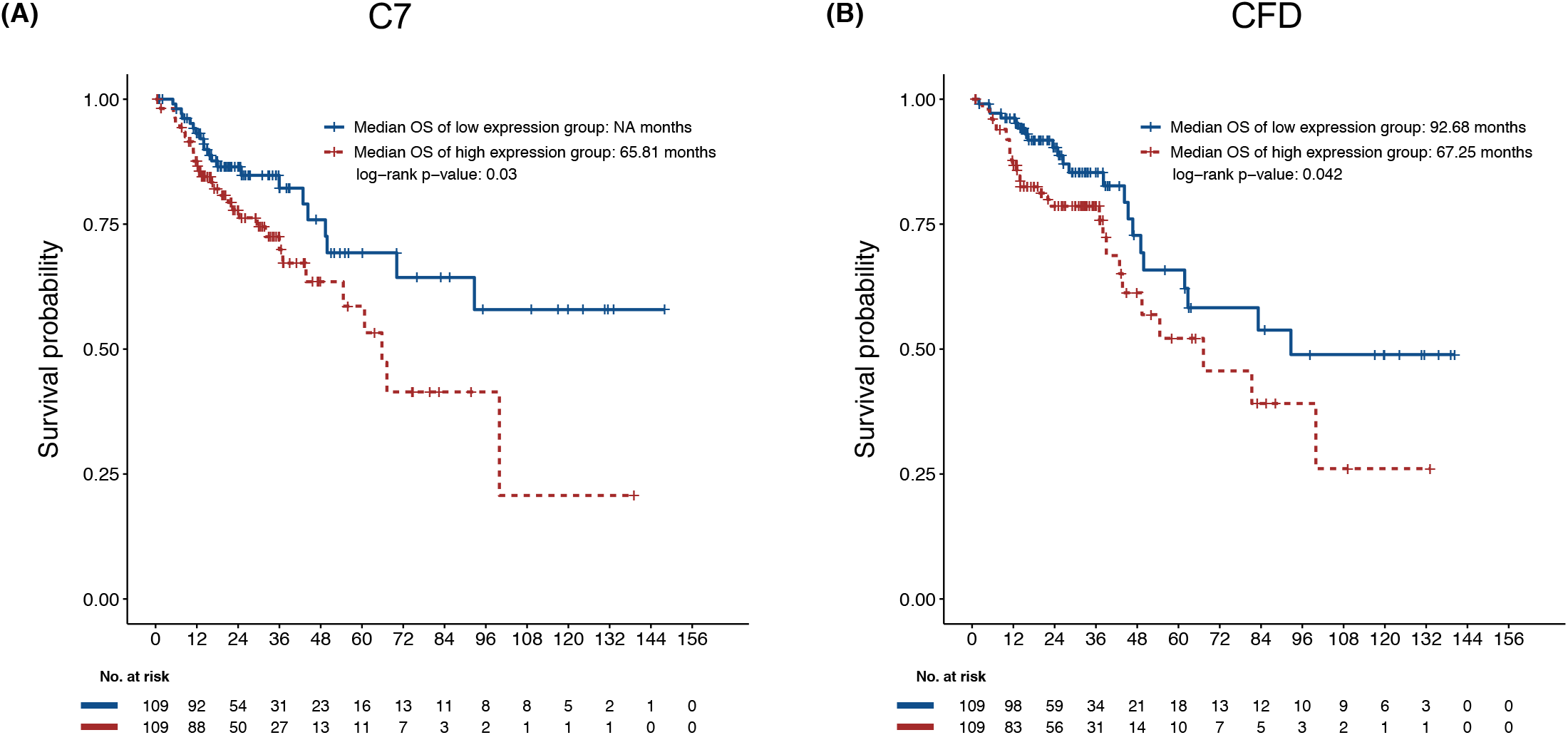
Kaplan-Meier curves of colorectal adenocarcinoma patients from The Cancer Genome Atlas (TCGA). Overall survival of TCGA colorectal adenocarcinoma patients in high (brown color line) and low (blue color line) expression groups of (A) C7 and (B) CFD. p-value was calculated by log-rank test. NA indicates that the median overall survival time is not reached. OS: overall survival.

**Supplemental Figure 4:**
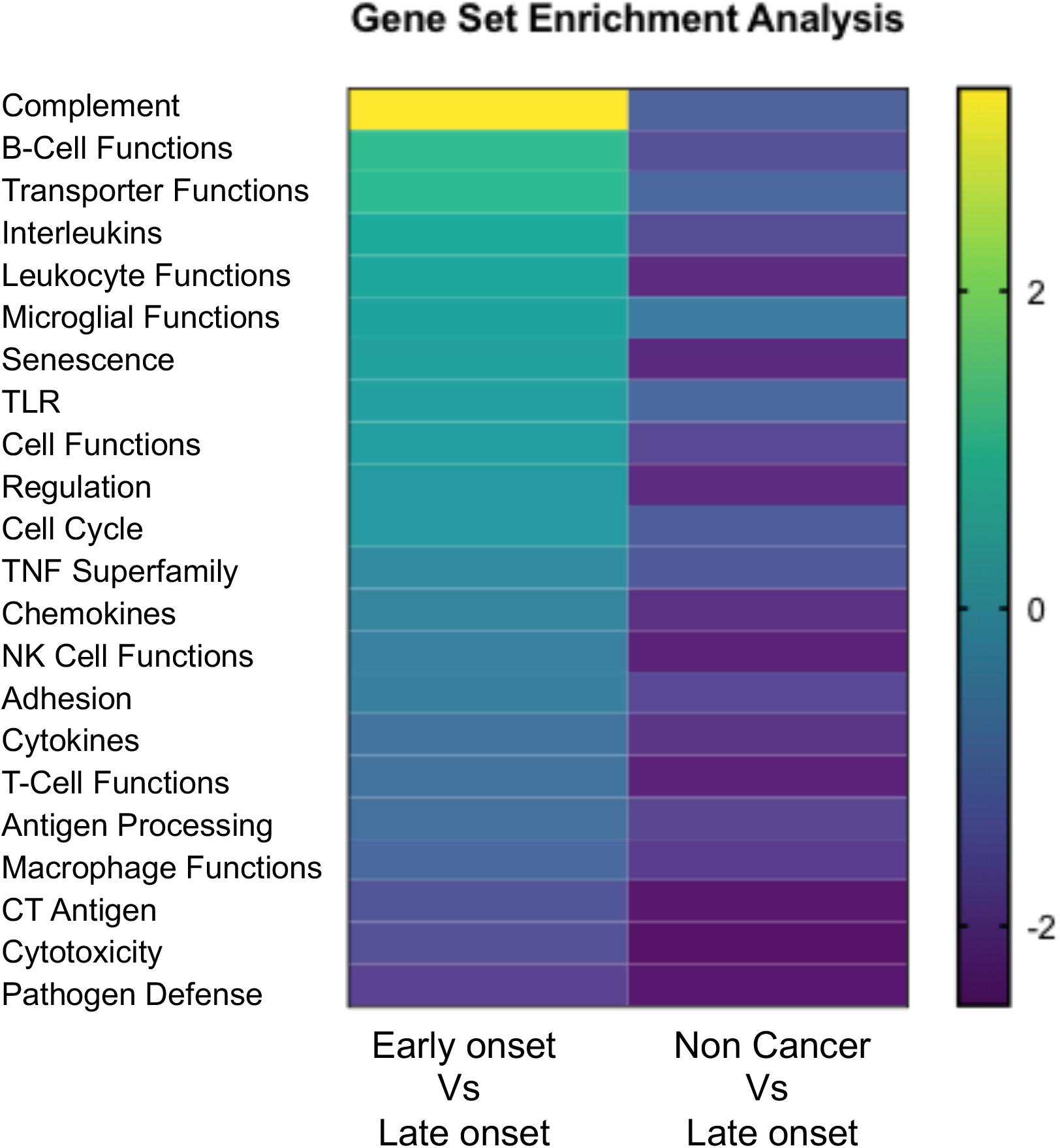
Heatmap of global significance scores for gene set enrichment between early and late onset colorectal cancer. The global significance score is calculated as the square root of the mean squared t-statistic for the genes in a gene set, with t-statistics coming from the linear regression underlying our differential expression analysis.

